# TICTAC: Target Illumination Clinical Trial Analytics with Cheminformatics

**DOI:** 10.1101/2025.01.23.25321034

**Authors:** Jeremiah I Abok, Jeremy S Edwards, Jeremy J Yang

## Abstract

**Introduction:** Identifying disease–target associations is a pivotal step in drug discovery, offering insights that guide the development and optimization of therapeutic interventions. Clinical trial data serves as a valuable source for inferring these associations. However, issues such as inconsistent data quality and limited interpretability pose significant challenges. To overcome these limitations, an integrated approach is required that consolidates evidence from diverse data sources to support the effective prioritization of biological targets for further research.

**Methods:** We developed a comprehensive data integration and visualization pipeline to infer and evaluate associations between diseases and known and potential drug targets. This pipeline integrates clinical trial data with standardized metadata, providing an analytical workflow that enables the exploration of diseases linked to specific drug targets as well as facilitating the discovery of drug targets associated with specific diseases. The pipeline employs robust aggregation techniques to consolidate multivariate evidence from multiple studies, leveraging harmonized datasets to ensure consistency and reliability. Disease–target associations are systematically ranked and filtered using a rational scoring framework that assigns confidence scores derived from aggregated statistical metrics.

**Results:** Our pipeline evaluates disease–target associations by linking protein-coding genes to diseases and incorporates a confidence assessment method based on aggregated evidence. Metrics such as meanRank scores are employed to prioritize associations, enabling researchers to focus on the most promising hypotheses. This systematic approach streamlines the identification and prioritization of biological targets, enhancing hypothesis generation and evidence-based decision-making.

**Discussion:** This innovative pipeline provides a scalable solution for hypothesis generation, scoring, and ranking in drug discovery. As an open-source tool, it is equipped with publicly available datasets and designed for ease of use by researchers. The platform empowers scientists to make data-driven decisions in the prioritization of biological targets, facilitating the discovery of novel therapeutic opportunities.

## 1 Introduction

ClinicalTrials.gov, managed by the U.S. National Library of Medicine (NLM), is a critical public repository that enhances transparency and accessibility in biomedical research. Established in 2000 under the Food and Drug Administration Modernization Act (FDAMA) of 1997, it serves as a centralized platform for registering clinical studies and disseminating trial results, addressing challenges related to data accessibility and reliability (Zarin et al., 2005; Tse et al., 2018). By supporting clinical trial registration, the database provides researchers, policymakers, and the public with comprehensive information about study designs, methodologies, and outcomes. Adherence to global reporting requirements, including those mandated by the International Committee of Medical Journal Editors (ICMJE) and the World Health Organization (WHO), ensures standardized practices across the research community (Laine et al., 2007). ClinicalTrials.gov aligns with FAIR (Findable, Accessible, Interoperable, and Reusable) principles (Wilkinson et al., 2016), employing standardized data formats to enhance interoperability across platforms and disciplines. It curates metadata, harmonizes submissions, and implements consistent reporting standards, addressing challenges such as incomplete reporting, variability in trial methodologies, and inconsistencies in diagnostic criteria (Riveros et al., 2013). The platform supports centralized trial registration and compliance with legal and ethical mandates for transparency (Zarin et al., 2011). Its study results database enables researchers to submit and access summary results, fostering evidence-based decision-making and mitigating publication bias through the inclusion of unpublished trial data (Prayle et al., 2012). Furthermore, integration with global registries, such as the WHO’s International Clinical Trials Registry Platform (ICTRP), promotes harmonization and accessibility of clinical research data worldwide. Despite its significant contributions, ClinicalTrials.gov faces challenges including variations in reporting quality, delays in result submissions, and inconsistencies in terminology (DeVito et al., 2020). The platform continues to evolve by introducing advanced data validation tools, promoting adherence to global reporting standards, and collaborating with stakeholders to refine metadata frameworks and bridge reporting gaps. As a robust and scalable platform for managing clinical trial data, ClinicalTrials.gov fosters transparency and enhances research reproducibility while empowering the scientific community and improving public trust.

The Database for Aggregate Analysis of ClinicalTrials.gov (AACT) was introduced to address challenges in analyzing aggregate data from ClinicalTrials.gov, such as inconsistent data structures, variability in nomenclature, and evolving data collection practices. Its purpose is to enhance the usability of ClinicalTrials.gov data by consolidating and normalizing information, enabling more effective aggregate analysis, policy studies, and systematic evaluations of clinical trial attributes and trends (Tasneem et al., 2012). AACT transforms raw clinical trial data into a structured, enriched, and analyzable format, integrating Medical Subject Headings (MeSH) terms and advanced curation techniques to ensure consistency and usability (Tasneem et al., 2012). By enabling systematic reviews, policy analysis, and diverse applications, AACT plays a pivotal role in evaluating global trial trends, aligning with FAIR principles, and driving innovation in clinical research. Despite challenges like reliance on MeSH hierarchies and limited global representation, ongoing advancements in ontology integration and interoperability aim to position AACT as an indispensable resource for the future of clinical trial analysis.

The core principle of pharmacology is that drugs, whether small molecules or biologics are designed to specifically interact with target molecules, often proteins, to modulate physiological processes and influence disease progression (Moffat et al., 2017; Scannell et al., 2012). Advanced methods in the pharmaceutical industry facilitate the discovery and optimization of these drugs, addressing challenges in efficacy, dosing, and safety for market approval (Hay et al., 2014). However, analysis of drug development pipelines reveals that insufficient efficacy, particularly in late-stage clinical trials, is a primary cause of failure, often due to inadequate validation of the target’s role in disease physiology (Ledford, 2011; Liu et al., 2021). This highlights the need for rigorous evidence supporting target-disease associations to improve success rates and minimize costly late-stage failures (Dahlin et al., 2015). Traditionally, drug targets were selected based on experimental evidence linking their modulation to disease outcomes (Muller & Milton, 2012). Recent advancements in high-throughput technologies, such as sequencing, genotyping, and mass spectrometry, have enhanced our ability to characterize biological samples, uncovering new opportunities to understand disease mechanisms (Vincent et al., 2015; Huang et al., 2011). Furthermore, the growing repository of clinical trial data, alongside extensive literature, serves as a valuable resource for identifying targets and generating hypotheses to inform the drug discovery process (Lysenko et al., 2018).

Here, we present TICTAC (Target Illumination Clinical Trial Analytics with Cheminformatics), an application designed to illuminate understudied drug targets by leveraging aggregated data from AACT. TICTAC enables ranking, filtering, and interpretation of inferred disease–target associations, assigning scores derived from aggregated evidence linking diseases to protein-coding genes mapped from drugs. This study outlines the analytical framework and interpretability of TICTAC, addressing statistical and semantic challenges. TICTAC demonstrates the application of data science in achieving scientific consensus and improving interpretability. The validation of disease-target associations is critical for ensuring the accuracy and reliability of biomedical datasets. This study also presents a methodology for validating the TICTAC disease-gene associations against MedlineGenomics (formerly Genetics Home Reference) by leveraging standardized disease terminologies, such as Disease Ontology IDs (DOIDs) (Schriml et al., 2022) and UMLS Concept Unique Identifiers (CUIs) (McInnes et al., 2007). By comparing disease-target associations across these datasets, the aim is to quantify overlap, identify areas of divergence, and provide insights into the consistency and reliability of the two data sources.

## 2 Material & Methods

### 2.1 AACT Data Preprocessing

The initial step in our methodology involved applying NextMove LeadMine (version 3.14.1). LeadMine, developed by NextMove Software, is a commercial text mining tool that identifies and annotates chemical entities, protein targets, genes, diseases, species, and more using curated grammars and dictionaries with advanced capabilities like correcting misspelled terms with CaffeineFix technology and supports chemical entity recognition in multiple languages, including Chinese and Japanese(Lowe & Sayle, 2015). This tool was utilized to analyze intervention names and study descriptions retrieved from ClinicalTrials.gov, enabling the identification of unique drug names and their corresponding SMILES (Simplified Molecular Input Line Entry System) representations. However, many terms encountered, such as "placebo," "test product," "medication," and "chemotherapy," lacked specific structural chemical information. For disease-named entity recognition (NER), we leveraged the JensenLab Tagger (Cook & Jensen, 2019). The JensenLab Tagger is a dictionary-based NER tool designed to identify and annotate entities such as genes, proteins, species, diseases, and other biomedical terms within text. It has been instrumental in text-mining applications, including extracting protein-protein interactions and annotating biomedical literature. The source code is available on GitHub: https://github.com/larsjuhljensen/tagger. This tool was applied to trial descriptions to identify and categorize diseases, resolving disease mentions to standardized terms in the Disease Ontology (DOID). This enhanced the consistency and granularity of disease data across sources. Subsequently, compound-target mapping was conducted to identify potential drug targets. Chemical entities were mapped to PubChem(Kim et al., 2021) using the PUG REST API with SMILES-based exact search and to ChEMBL(Zdrazil et al., 2024) using REST API queries via InChIKey. Biological targets were then mapped from ChEMBL bioassays and linked to the Integrated Disease-Target Knowledgebase (IDG-TCRD/Pharos)(Sheils et al., 2021) using UniProt IDs (The UniProt Consortium, 2023). This systematic mapping established relationships between chemical entities, their biological targets, and associated diseases, forming a foundation for data aggregation efforts.

Our dataset, extracted from the Aggregate Analysis of ClinicalTrials.gov (AACT) database as of September 30, 2024, comprised 507,584 studies, each identified by a unique NCT_ID. These studies referenced 901,776 publications, associated with 632,153 PubMed IDs (PMIDs) and 127,455 RESULT-type references, spanning 170,697 unique NCT_IDs. For drug-related data, 133,760 unique drug names were linked to 365,878 unique intervention IDs. It is worth noting that individual NCT_IDs often referenced multiple drugs, with synonymous naming conventions contributing to challenges in precise drug identification.

Our analysis focused exclusively on interventional drug studies, excluding observational studies. Of the total dataset, 388,958 (76.6%) studies were classified as interventional, with 177,780 designated as interventional drug studies, each linked to a unique NCT_ID. To refine the chemical data, NextMove LeadMine was used to resolve drug names into standardized chemical structures via SMILES notation. This process identified 6,595 unique SMILES associated with 23,982 unique intervention IDs and 19,662 unique drug names. The resulting dataset included 901,776 study references and 632,153 PubMed IDs, offering a robust basis for linking identified entities in clinical trial data to biomedical literature.

### 2.2 Computations for Disease-Target Associations

#### nStudyNewness

The **nStudyNewness** metric quantifies the recency-weighted relevance of clinical studies associated with each disease-gene pair, reflecting the higher impact of newer studies. This is achieved using an exponential decay function that prioritizes recent studies.

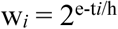

where:

- *t_i_* is the age of the study in years,
- h is the half-life determining the decay rate (h = 5 years for studies ≤ 10 years old and h=10 years for older studies).

The total nStudyNewness for a disease-target pair is:

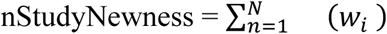

where *N* is the total number of studies associated with the disease-target pair.

#### nPublicationWeighted

The **nPublicationWeighted** metric assigns weights to publications based on their reference type, accounting for their varying levels of impact on disease-target associations. For each publication *i* with reference type *r_i_*, the weight *w_i_* is defined as:

- *w_i_*=1.0: If the publication reference type *r_i_* is a **Result** (denoted by 0.0), it is given the highest weight.
- *w_i_*=0.5: If *r_i_* is a **Background** reference (denoted by 1.0), it is assigned a medium weight.
- *w_i_*=0.25: If *r_i_* is a **Derived** reference (denoted by 2.0), it is given the lowest weight.

#### Ranking Associations

To facilitate the prioritization of disease-gene associations, rankings are computed based on multiple metrics, culminating in an overall score.

1. **Rank Computation:** For each metric (nDiseases, nDrug, nStud, nPub, nStudyNewness, nPublicationWeighted), the rank *R_m,j_* for disease-target pair *j* is computed in descending order:

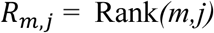

2. **Mean Rank:** The average rank across all metrics for a disease-target pair *j* is:

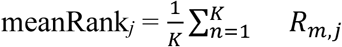

where *K* is the total number of metrics.

3. **Percentile Rank:** Percentile rank is calculated to normalize the meanRank values:

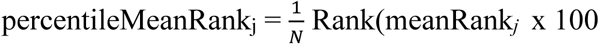

where *N* is the total number of disease-target pairs.

4. **Mean Rank Score:** The final mean rank score, providing a scale from 0 to 100, is:

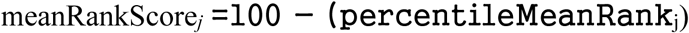

This methodology ensures inferred disease-target associations are systematically ranked based on multiple evidence metrics, with the highest meanRankScore indicating the most promising targets for further research and development.

### 2.3 Validation Method

The validation methodology included key steps to map and compare disease-target associations between TICTAC and MedlinePlus Genetics (MedlineGenomics) (U.S. National Library of Medicine, n.d). The mapping from Disease Ontology was processed to link DOIDs with UMLS CUIs and validate data integrity through unique counts. MedlinePlus genetics conditions were formatted to align with these mappings, ensuring compatibility and accuracy. TICTAC disease-target associations were similarly prepared by standardizing identifiers, refining data fields, and verifying counts of key entities. Common CUIs between datasets were identified, and their corresponding associations were extracted for comparison. Overlaps between the datasets were evaluated by analyzing shared associations, with the percentage of MedlinePlus Genetics associations present in TICTAC calculated. The validation assessed dataset integrity, overlap in conditions (common CUIs), and overlap in disease-target associations to measure alignment and reliability.

## 3 Results

### 3.1 The TICTAC Application

TICTAC supports drug target identification by scoring and ranking associations between drug targets (protein-coding genes) and diseases. The TICTAC workflow aggregates and filters inferred clinical trial findings to generate actionable insights. These insights can be leveraged to enhance target prioritization through interactive visualizations and hit lists (Fig. 2), enabling users to identify the strongest, evidence-supported associations.

**Fig. 1.**
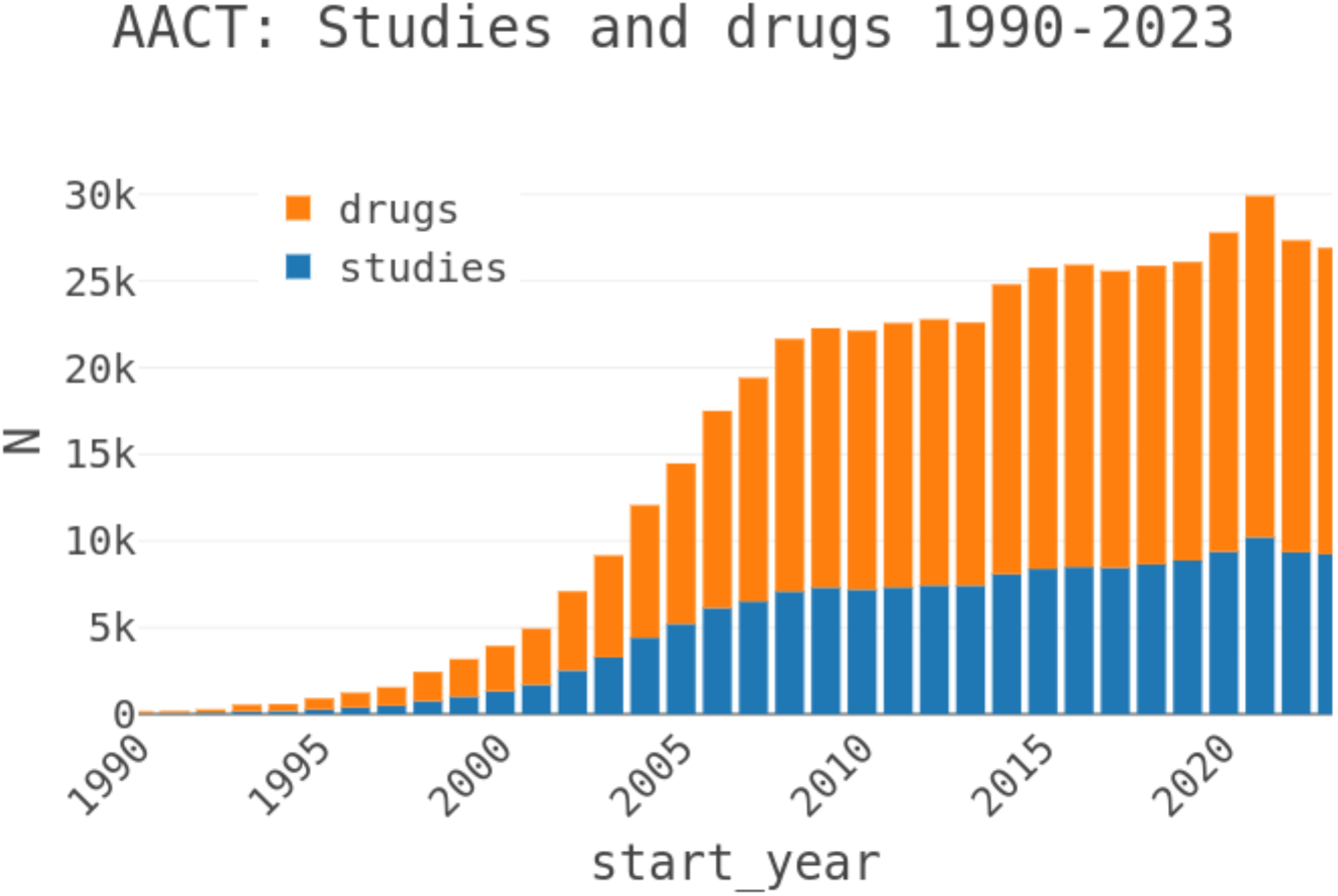
Clinical Trial study counts by year indicating growth and trends

**Fig. 2.**
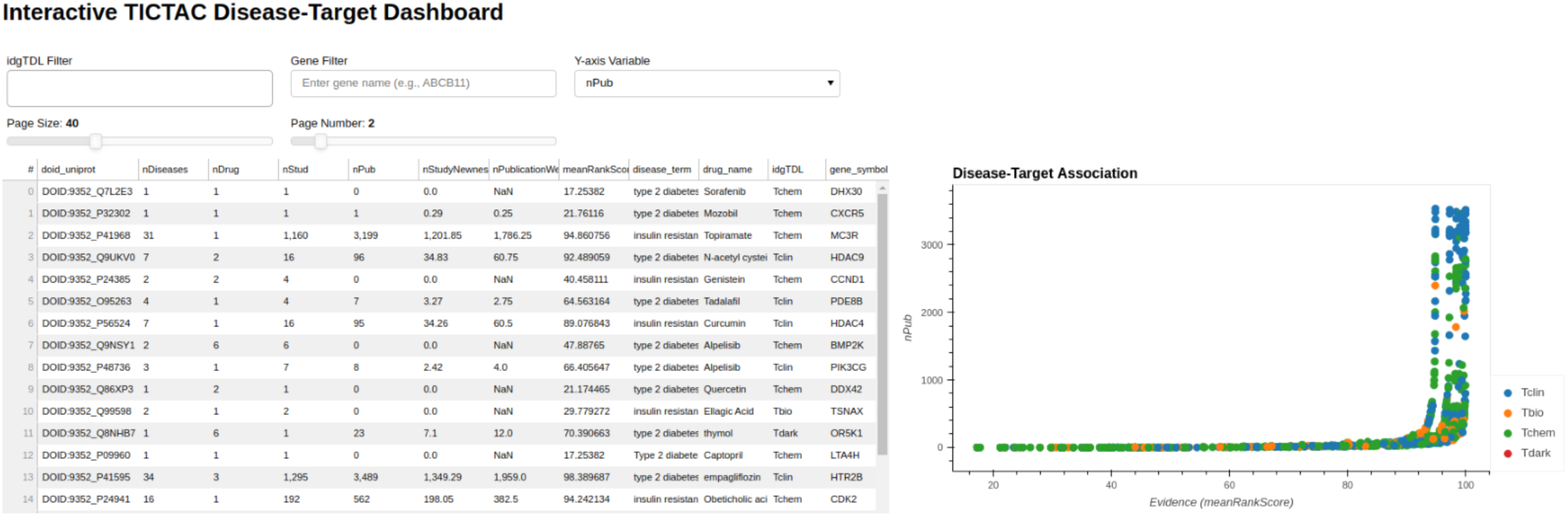
TICTAC dashboard. The image displays genes currently associated with the disease ’type 2 diabetes/insulin resistance’ (DOID:9352)

Hits in the TICTAC dashboard are ranked by **meanRankScore**, as described in Section 2. The scatterplot presents evidence (meanRankScore) on the X-axis versus publication count (nPub) on the Y-axis, visually representing disease-target associations. The workflow allows users to filter results using query parameters such as disease terms (e.g., "type 2 diabetes/insulin resistance," DOID:9352) and gene symbols. Data points in the scatterplot are colored by target development level (TDL), a knowledge-based classification system that categorizes human proteins into four distinct groups (Oprea et al., 2018) as Tclin (Santos et al., 2017), Tchem, Tbio, and Tdark for the comprehensiveness of exploration from clinical, chemical, and biological perspectives. This framework enables the prioritization of disease-target pairs based on their evidence levels and functional classifications, facilitating drug discovery efforts and identifying novel targets for further research.

### 3.2 Agreement-Based Validation of TICTAC and MedlinePlus Genetics Datasets

The agreement-based validation process revealed that the TICTAC dataset includes 2,243 unique Disease Ontology IDs (DOIDs) and 2,022 unique gene symbols, while the MedlinePlus Genetics dataset contains 1,216 conditions mapped to Concept Unique Identifiers (CUIs) and 2,142 unique CUIs. A total of 193 CUIs were shared between the two datasets, allowing for a comparative analysis of associations. For the shared CUIs, the TICTAC dataset encompassed 63,569 associations involving 1,804 gene symbols, whereas the MedlinePlus Genetics dataset comprised 1,247 associations involving 967 gene symbols. Notably, 136 associations overlapped between the two datasets, accounting for 10.91% of the MedlinePlus Genetics associations. This indicates that the TICTAC dataset contains a high proportion of novel associations, with an estimated 89% of associations being unique to TICTAC. These results highlight the potential of the TICTAC dataset to contribute significant novel insights, while also demonstrating consistency between the datasets for a subset of shared associations. This agreement-based approach underscores the complementary nature of these resources and the value of TICTAC for expanding the landscape of disease-target associations for further investigation.

### 3.3 Using TICTAC for Drug Target Illumination

#### 3.3.1 Exploring Understudied Genes in Type2 Diabetes/Insulin Resistance: Integrating Metrics for Target Discovery Potential

Fig. 3 highlights examples of understudied (Tbio) genes associated with "type 2 diabetes/insulin resistance" within the TICTAC framework. The analysis focuses on genes classified as Tbio, indicating they are less characterized and have limited research attention compared to more ‘established targets’. The table provides key metrics for each gene-disease pair, including the number of diseases (nDiseases), drugs (nDrug), studies (nStud), and publications (nPub), along with evidence prioritization scores such as nStudyNewness and meanRankScore. These metrics collectively illustrate the relative evidence strength and research activity for each gene-disease pair. The analysis reveals significant variability in the evidence supporting these genes. For instance, some genes, such as UGT1A10 (41 publications, meanRankScore = 97.87784), have substantial supporting evidence, while others, such as COQ8A (0 publications, meanRankScore = 46.33545), remain less explored in the literature. Importantly, all listed genes have some level of therapeutic association, as reflected in the nDrug column, which indicates the number of drugs linked to each gene. For example, MAP4K4 and RAC1 are linked to drugs such as Sorafenib and Dasatinib, while other genes, such as CISD2, are associated with broader therapeutic contexts. The nStudyNewness metric emphasizes the relevance of recent evidence for these gene-disease pairs, with genes like CISD2 demonstrating strong support from newer studies and achieving a high meanRankScore (98.92213). Such metrics underscore the potential for these Tbio genes to serve as new therapeutic targets for metabolic disorders. Genes with high meanRankScore and robust evidence, such as CISD2, may warrant further investigation in drug discovery efforts. Conversely, genes with minimal publications, such as COQ8A, represent opportunities for expanding research into their roles in type 2 diabetes and insulin resistance.

**Fig. 3.**
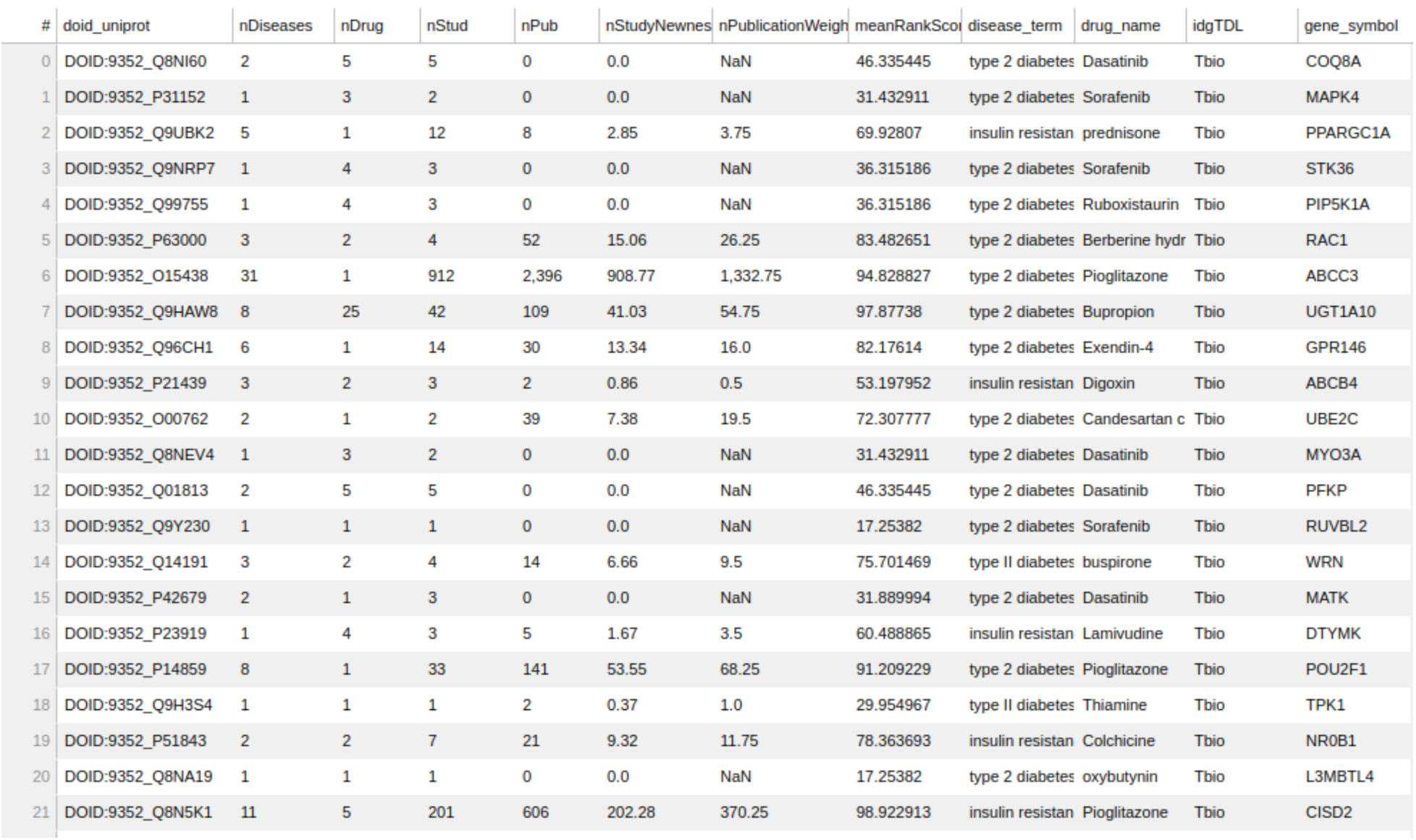
Examples of understudied (Tbio) genes for disease ’type 2 diabetes/insulin resistance’ in TICTAC

Overall, this analysis of Tbio genes in the context of type 2 diabetes/insulin resistance highlights the utility of TICTAC in identifying and prioritizing understudied targets for further research and therapeutic development. By integrating metrics such as meanRankScore, nPub, and nStudyNewness, the framework enables systematic exploration of gene-disease associations, providing actionable insights for drug discovery.

#### 3.3.2 Integrating Evidence for Disease-Target Associations: Insights from the TICTAC Provenance Dashboard

Figure 4 presents the TICTAC Provenance Dashboard, which provides detailed reference data supporting the association between the MCR4 (Melanocortin receptor 4) gene and type 2 diabetes/insulin resistance (DOID:9352). This dashboard aggregates references from clinical trials and publications, offering comprehensive insights into the evidence linking the gene to the disease. The framework for the provenance of this association includes several key elements. The **nct_id** column lists unique identifiers for clinical trials, while the **reference_type** column categorizes evidence as BACKGROUND (contextual information), RESULT (direct findings), or DERIVED (secondary findings). The **pmid** column includes PubMed IDs linking to corresponding publications, and the **citation** column provides bibliographic details such as titles, authors, journals, and publication years. The results reveal a mix of evidence types. BACKGROUND references highlight physiological mechanisms like insulin sensitivity and glucose metabolism foundational to type 2 diabetes research. RESULT references contribute direct findings, such as variability in cyclooxygenase inhibition in aspirin studies, while DERIVED references explore broader evidence, such as anti-psychotropic medication usage. For example, Larsen et al. (2012) offer insights into diabetes mechanisms.

**Fig. 4.**
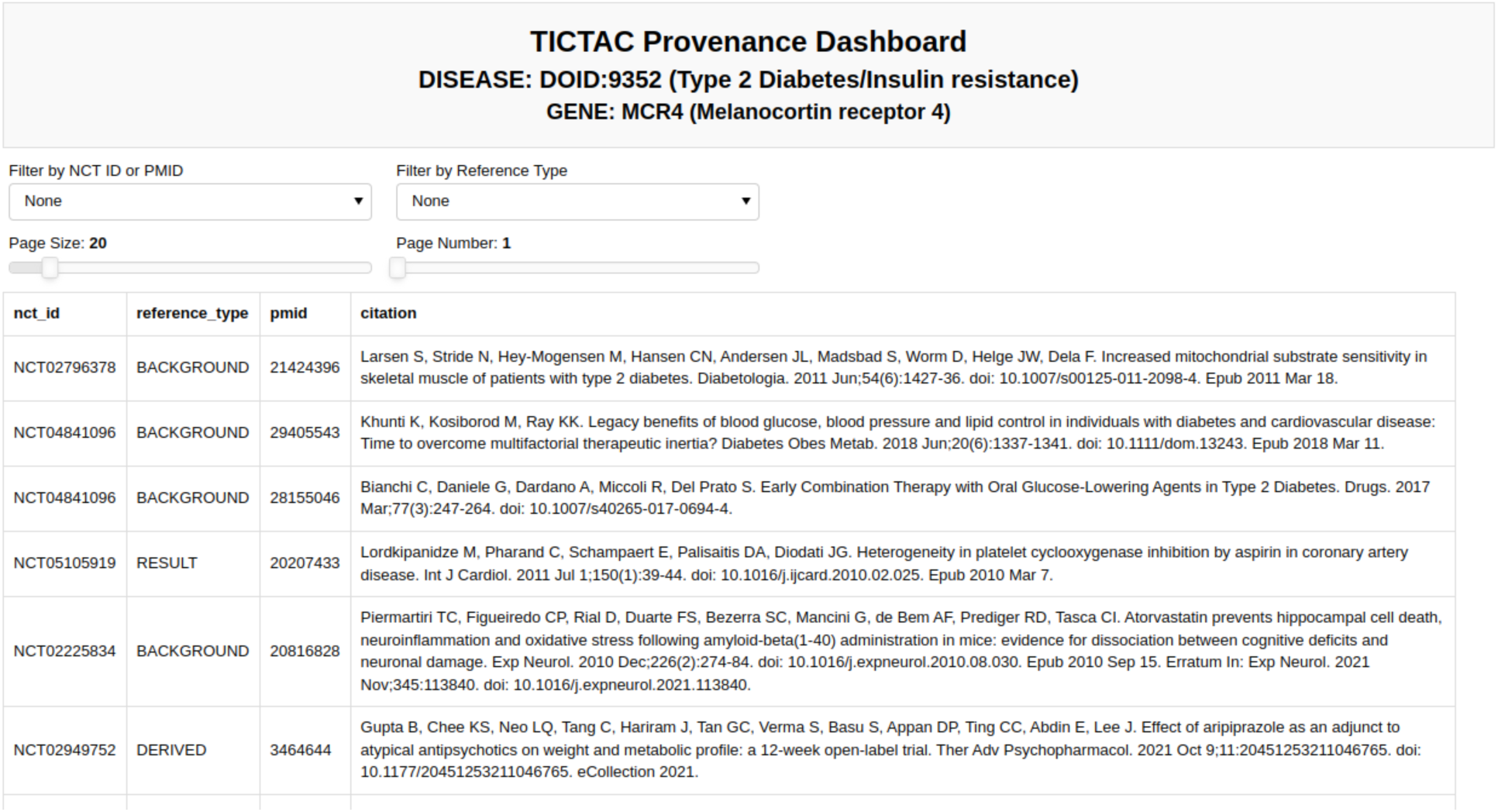
Provenance for association between gene MCR4 (Melanocortin receptor 4) and disease ’ type 2 diabetes/insulin resistance’.

Sodium-glucose cotransporter 2 (SGLT2) inhibitors have emerged as a novel therapeutic approach for type 2 diabetes, particularly in improving cardiovascular outcomes. These inhibitors reduce myocardial glucose uptake by up to 57%, inducing metabolic shifts toward fatty acid utilization (Lauritsen et al., 2021). Additionally, they lower myocardial blood flow (MBF) by approximately 13%, potentially due to diuretic effects that reduce blood pressure and alter renal hemodynamics (Kimura, 2016). While SGLT2 inhibitors significantly mitigate glucose uptake and blood flow, their broader cardioprotective effects likely involve mechanisms beyond these metrics. For example, they do not notably alter myocardial oxygen consumption, suggesting additional pathways are at play (Lauritsen et al., 2021; Huang et al., 2023). Despite these complexities, improved cardiac function and reduced heart failure risk highlight their therapeutic potential (Huang et al., 2023). A strong hypothesis thus emerges: SGLT2 inhibition improves cardiac outcomes in individuals with type 2 diabetes by inducing metabolic shifts, reducing myocardial blood flow, and enhancing cardiac efficiency. This framework underscores the importance of integrating diverse evidence types to establish robust disease-target associations. Direct access to clinical trial identifiers and PubMed references enhances reproducibility and credibility, making the TICTAC Provenance Dashboard a valuable resource for researchers exploring therapeutic targets in type 2 diabetes.

#### 3.3.3 Mapping Disease-Target Associations and Evidence in Lung Cancer Research

Figures 5 and 6 provide a comprehensive view of gene-disease associations related to lung cancer (DOID:1324), focusing on potential gene targets and the provenance of specific evidence linking the PGR (Progesterone receptor, UniProt: P06401) gene to the disease.

**Fig. 5.**
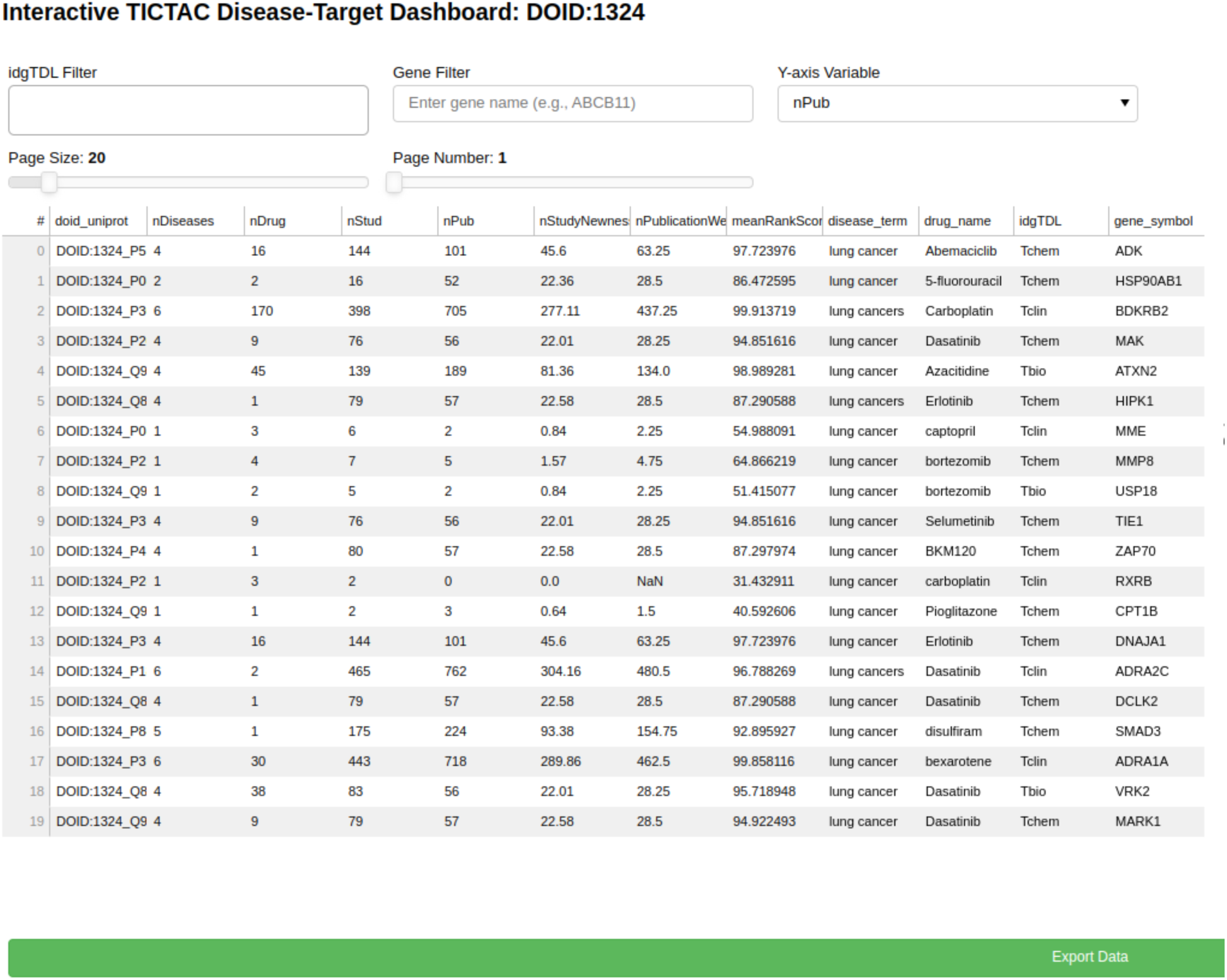
TICTAC dashboard, displaying a plot of genes associated with disease ’lung cancer’ (DOID:1324)

Figure 5 presents the TICTAC Disease-Target Dashboard, which highlights key metrics for genes associated with lung cancer. The dashboard includes metrics such as the number of diseases (nDiseases), drugs (nDrug), studies (nStud), and publications (nPub) related to each gene-disease pair. Additional evidence metrics such as nStudyNewness, which prioritizes recent studies, and PublicationWeight, which accounts for the relevance of evidence, are also shown. The meanRankScore provides a composite metric ranking associations based on the strength and quality of evidence. For each gene, the dashboard also lists the associated drugs and Target Development Level (TDL), a classification reflecting the druggability of the gene.Notable findings include highly ranked genes such as ADK (meanRankScore = 97.73276) and BDKRB2 (meanRankScore = 95.97179), which are supported by robust evidence from multiple studies, publications, and associated drugs. Therapeutically, drugs such as Abemaciclib, 5-Fluorouracil, and Carboplatin are frequently linked to the listed genes, emphasizing their relevance to lung cancer treatment. The genes in this dashboard are classified as Tchem, indicating their known potential as druggable targets, making this dashboard a critical resource for prioritizing genes with strong evidence for further exploration in lung cancer research and therapeutic development.

Figure 6 provides detailed evidence provenance for the association between the PGR (Progesterone receptor) gene and lung cancer. The TICTAC Provenance Dashboard aggregates references from clinical trials and publications to trace the supporting evidence. It includes clinical trial identifiers (nct_id), 2978 reference types categorized as RESULT for direct findings or BACKGROUND for supporting evidence, PubMed IDs (pmid), and detailed citations for each referenced study. The provenance dashboard reveals that the association between PGR and lung cancer is supported by both direct results and background studies. These references detail the role of therapies such as carboplatin in treating lung cancer, providing clinical context for PGR’s relevance. This detailed provenance ensures the traceability of the data and supports further exploration of PGR in lung cancer research.

**Fig. 6.**
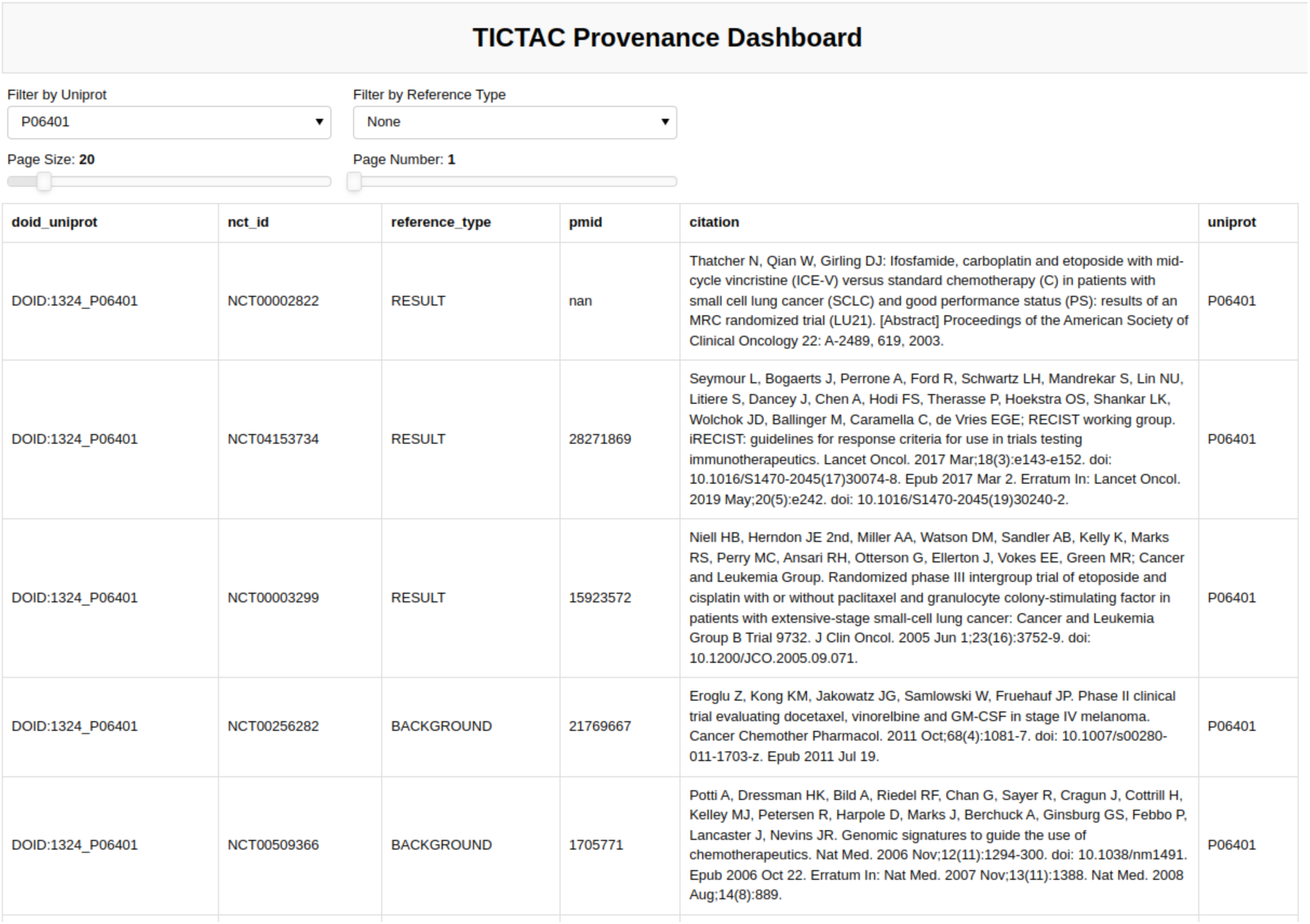
Provenance for association between **PGR** gene (Uniprot: P06401) ’ **Progesterone receptor**’ and disease ’lung cancer’

Taken together, Figures 5 and 6 illustrate the utility of TICTAC in mapping disease-target associations and integrating comprehensive evidence provenance. The TICTAC Disease-Target Dashboard enables the identification of high-priority gene targets and associated drugs, while the Provenance Dashboard provides detailed and traceable evidence for specific genes like PGR. This combination of tools allows for a robust prioritization framework, supporting translational research and advancing the discovery of potential therapeutic targets.

## 4 Discussion

### 4.1 Illuminating Knowledge Gaps in Targets

The National Institutes of Health’s (NIH) Illuminating the Druggable Genome (IDG) program is dedicated to advancing our understanding of understudied proteins within pivotal druggable families, including G-protein-coupled receptors (GPCRs), ion channels, and protein kinases (National Institutes of Health, 2024). By elucidating the roles of these proteins in health and disease, the program seeks to identify novel therapeutic targets and foster innovative drug development strategies.

For a detailed exploration of the objectives and methodologies of the IDG program, refer to "Unexplored therapeutic opportunities in the human genome" by Oprea et al. (Oprea et al., 2018). TICTAC aligns seamlessly with this mission, focusing on evaluating clinical trial evidence to reveal disease–target associations. Unlike other platforms, such as Open Targets (Ghoussaini et al., 2021; Ochoa et al., 2021), which leverage a blend of data, supervised machine learning, and external sources, TICTAC strictly aggregates evidence from the AACT database. This approach minimizes biases inherent in curated training data and domain-specific assumptions, offering interpretable results that are grounded in experimental provenance and reproducible methodologies. By integrating its automated, sustainable workflow into resources such as the Pharos portal (Nguyen et al., 2017; Sheils et al., 2021), TICTAC complements existing tools for target illumination. While Open Targets supports scientists by enhancing associations with external validation, TICTAC prioritizes the rigor of direct clinical trial data. This makes it particularly suited for downstream users who require traceable, transparent insights into disease–target relationships.

### 4.2 Transforming Data into Actionable Knowledge

In genomics and drug discovery, where data overload is a persistent challenge, transforming raw information into actionable knowledge remains critical. Specialized tools aid in uncovering insights, but the interpretation and integration of these findings often demand expertise that non-specialists may lack. TICTAC addresses this gap by introducing a layer of abstraction that is not only accessible but also directly aligned with the objectives of drug discovery scientists. TICTAC’s design philosophy rests on simplicity and scientific axioms, such as prioritizing evidence from independent confirmatory results (Yang et al., 2021). This emphasis ensures the interpretability of findings and highlights clinical trial results as a cornerstone of evidence-based reasoning. While correlation does not equate to causation, it builds a foundation of plausibility, fostering hypothesis generation and prioritization. With this context in mind, TICTAC provides a pragmatic, rational framework for ranking research hypotheses. Clinical trials are often influenced by experimental noise and systematic uncertainties stemming from factors like the COVID-19 pandemic’s impact on follow-ups (Servick, 2020), incomplete data from missing participants (Verzilli & Carpenter, 2002), and biases in endpoint evaluations (Chen et al., 2020). Ambiguities in defining study populations and inadequate reporting of sample sizes further complicate result interpretation (Frampton & Shepherd, 2008). Addressing these challenges through improved methodologies and reporting standards is essential to enhance the validity and reliability of trial outcomes. While acknowledging that experimental noise and systematic uncertainties can accompany clinical trial data, the approach ensures that aggregated insights are meaningful and usable for non-specialist stakeholders in drug development.

### 4.3 Designing for Seamless Integration and Confidence

Biomedical knowledge discovery thrives on the integration of diverse, heterogeneous data sources, reflecting the inherent complexity of the field. However, challenges related to provenance, interpretability, and confidence frequently undermine these efforts. TICTAC addresses these concerns by employing simple yet robust metrics, such as unbiased meanRank scores, to evaluate and rank disease–target associations. One of TICTAC’s key innovations lies in its transparent approach to confidence assessment. By restricting provenance to the linked publications, the platform ensures enhanced interpretability and traceability. This minimalist yet methodical approach reduces the accumulation of errors and confidence decay that often plague systems integrating multiple, heterogeneous data sources. Continuous confidence scores allow for dynamic algorithmic weighting and filtering, supporting downstream applications that require both precision and flexibility. The clear use of standardized identifiers and semantics strengthens integration across biomedical resources, paving the way for consistent cross-platform compatibility. Figure 7 illustrates TICTAC’s workflow and interfaces, underscoring its defined role in disease–target discovery pipelines. With its focus on transparency, interpretability, and confidence, TICTAC offers a practical and scientifically rigorous tool for advancing biomedical research and drug discovery.

**Fig 7:**
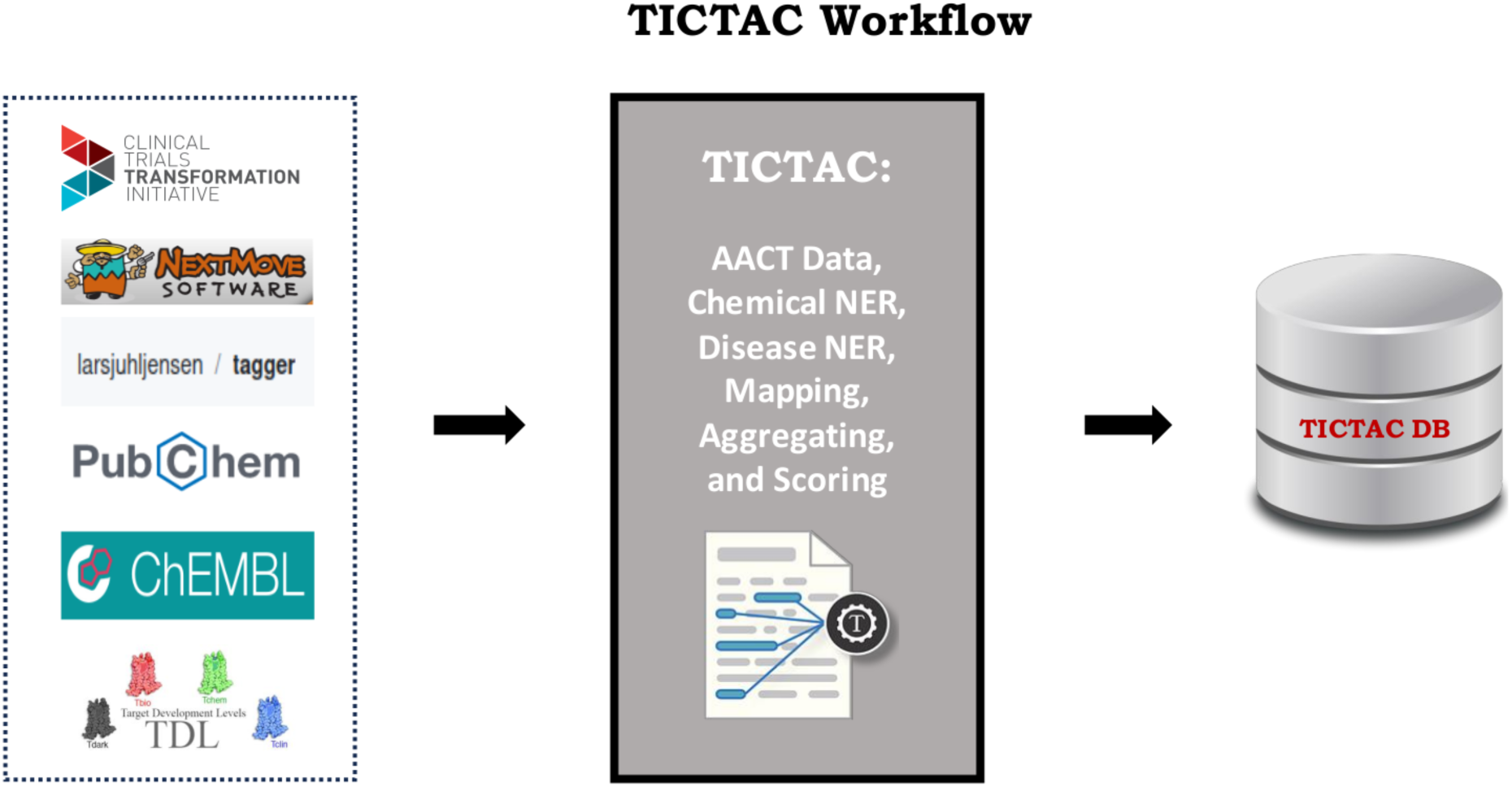
TICTAC data sources and interfaces. TICTAC integrates clinical trial data from the AACT db and cross referenced other sources to rank disease–target associations. These associations can be accessed through the TICTAC github repository.

### 4.4 Validation

The validation process revealed a 10.91% overlap between MedlineGenomics and TICTAC disease-gene associations for the 193 shared CUIs, indicating some level of consistency while also highlighting significant differences between the datasets. These discrepancies arise from variations in data collection methods, terminological standards, and the granularity of gene-disease associations. The low percentage of overlap underscores the need for improved standardization in disease-gene datasets. This process emphasized the value of standardized identifiers such as DOIDs and CUIs for aligning and comparing biomedical data. It also highlighted potential gaps in data completeness or detail, which could affect the utility of these datasets in downstream research, and reinforced the importance of integrating multiple sources to achieve a comprehensive understanding of disease-gene relationships.

## 5 Conclusion

The conventional "one gene, one function, one trait" paradigm, as critiqued by Visscher et al. (2017), is no longer adequate for understanding the intricate mechanisms underlying diseases. This shift in understanding highlights the need for advanced tools that can navigate the multifaceted nature of genetic and clinical data, enabling researchers to uncover meaningful relationships between genes, traits, and diseases (Yang et al., 2021). Modern biomedical research demands tools that move beyond oversimplifications to provide actionable insights grounded in real-world data. TICTAC rises to this challenge by offering a dynamic platform tailored for drug target hypothesis generation and refinement, leveraging clinical trial data and metadata to bridge the gap between data complexity and practical application. Aligned with the increasing focus on interpretable machine learning and explainable artificial intelligence (XAI) (Adadi & Berrada, 2018), TICTAC employs transparent, evidence-driven methodologies to elucidate disease–target associations. Its foundation rests solely on clinical trial data and metadata, supported by rational, intuitive evidence metrics, and underpinned by a robust open-source pipeline designed for continuous improvement and scalability. TICTAC’s adaptability allows it to function either as a standalone resource or as a component integrated with other analytical interfaces. By simplifying the exploration of complex clinical data, TICTAC contributes meaningfully to the identification and prioritization of drug targets, offering a practical and evolving tool for advancing biomedical research and translational science.

## Data Availability

All data produced are available online at https://github.com/unmtransinfo/TICTAC, as described in the manuscript, or, available upon reasonable request to the authors.

## 7 Author Contributions

## 12 Supplementary Material

## 13 Data Availability Statement

The datasets generated and analyzed for this study as well as the source code can be found in the TICTAC repository (https://github.com/unmtransinfo/TICTAC).

